# The impact of patient ethnicity on cancer incidence following platelet count and C-reactive protein tests in English primary care: a cohort study of 5 million patients

**DOI:** 10.64898/2026.03.03.26347503

**Authors:** Melissa Barlow, Liz Down, Luke T A Mounce, Samuel W D Merriel, Jessica Watson, Tanimola Martins, Sarah E R Bailey

## Abstract

**Background:** Platelet count and C-reactive protein (CRP) are blood tests commonly used in primary care as part of diagnostic work up for symptomatic patients. Abnormal results of these tests can indicate an undetected cancer; however, it is not known whether the association between an abnormal test result and cancer risk varies by patient ethnicity.

**Methods:** This cohort study used routinely collected primary and secondary health care records in England with linkage to national cancer registry data. Included patients had a record of ethnicity, no prior malignancy, a platelet count or CRP record between 1^st^ January 2010 and 31^st^ December 2017, and were aged 40 years or over at the time of that test. Ethnicity was categorised as White, Asian, Black, Other, and Mixed. Multi-level logistic regression models estimated cancer incidence within one-year of testing, adjusted for age, sex, comorbidities, BMI, deprivation, and year of test.

**Results:** Among 4,948,342 patients with a platelet record and 811,559 with a CRP record, one-year cancer incidence was highest among White patients and lowest among Asian patients. Following a normal platelet count, cancer incidence was 1.3% (95% CI 1.3-1.3%) for White patients and 0.63% (0.60-0.66%) for Asian patients; following thrombocytosis, incidence increased to 4.1% (4.0-4.2%) and 1.8% (1.5-2.0%), respectively. After a normal CRP result, cancer incidence was 1.5% (1.4-1.5%) for White patients and 0.79% (0.71-0.88%) for Asian patients, rising to 3.6% (3.5-3.7%) and 1.9% (1.7-2.2%) for a high CRP result, respectively. No significant interactions were found between ethnicity, blood test result, and overall cancer diagnosis, and similar diagnostic odds ratios (dOR) were observed across all ethnic groups.

However, for colorectal cancer, Black patients with abnormal results showed higher diagnostic odds ratios (dOR) compared with White patients, relative to a normal result. The dOR for thrombocytosis was 11.1 (7.8-15.6) for Black patients versus 5.7 (5.4-6.0) for White patients (interaction *p*-value <0.001), and for raised CRP was 4.1 (2.6-6.6) for Black patients versus 2.5 (2.3-2.7) for White patients (interaction *p*-value=0.043).

**Conclusion:** This large primary care study underscores the need for ethnically diverse cohorts when evaluating diagnostic tests to avoid widening healthcare inequalities.

## Introduction

The National Cancer Plan for England aims to prioritise earlier diagnosis, as well as taking action to reduce inequalities in cancer care (1). Inequalities in cancer diagnosis are consistently reported for Asian and Black communities in the UK (2–5). Despite generally experiencing lower incidence rates (6), patients from Asian and Black ethnic groups are more likely to be diagnosed at an advanced stage for some cancer types (2), experience longer time to diagnosis (3), and report worse experiences of cancer care than patients from White ethnic backgrounds (4,5). The factors driving these inequalities, particularly the role of primary care in the diagnostic pathways, are not fully understood.

Primary care remains the principal entry point for most cancer diagnostic journeys in the UK and substantial research has improving early diagnosis through this setting (7). Thrombocytosis (platelet count ≥400×10^9^/L) has a positive predictive value for undiagnosed cancer of 6% in women and 11% in men (8–11). and English national guidance recommends primary care clinicians to consider lung, colorectal, oesophageal-gastric, or uterine cancer in a patient with thrombocytosis (12). Platelet count is known to rise in response to infections and inflammation (13), and C-reactive protein (CRP), another inflammatory marker, has a positive predictive value of >3% in the detection of cancer in primary care (14).

However, studies of platelet count, CRP, and cancer risk have not examined the potential influence of ethnicity on these associations, despite evidence suggesting variation in their distribution across ethnic groups (8,9,11,14). A recent systematic review reported that Black patients from a healthy or general population in the USA were more likely to have higher platelet counts and higher CRP levels than their White or Asian counterparts. In contrast, studies in the UK and elsewhere reported no meaningful differences in platelet count by ethnicity, and no UK studies have yet described CRP distributions by ethnic group. Furthermore, it remains unclear whether these blood tests perform equally well in predicting cancer incidence across ethnic groups (15).

Evidence from studies of specific cancer markers (prostate specific antigen and cancer antigen-125) have shown variation in both baseline differences in test values by ethnicity in individuals without cancer, (16–19) and variation in test performance for cancer detection in primary care (20,21). The reasons for these differences are complex and not fully understood.

This study aims to 1) describe the distribution of platelet count and CRP levels for patients from different ethnic groups presenting to primary care in England, 2) assess cancer incidence following a blood test showing platelet count and CRP levels for patients from different ethnic groups in English primary care and 3) analyse the association of blood test result and cancer stage at diagnosis for different ethnicities.

## Methods

### Data sources

This cohort study used electronic English primary care records from the Clinical Practice Research Datalink (CPRD) Aurum dataset (22), linked to the Hospital Episode Statistics Admitted Patient Care (HES APC) dataset (23) and the National Cancer Registration and Analysis Service (NCRAS) cancer registry (24).

### Patient selection criteria

Patients were included if they had an ethnicity record in either the CPRD or Hospital Episode Statistics datasets, no prior history of malignancy (other than non-melanoma skin cancer), a platelet or CRP record between 1^st^ January 2010 and 31^st^ December 2017, and were aged 40 years or over at the time of that test.

### Outcome variables

Cancer diagnoses were identified from the cancer registry, and patients were classified as being diagnosed with cancer if a cancer diagnosis was recorded in the year following the date of their blood test. Patients with no cancer recorded in this timeframe (one-year) were classified as not having been diagnosed with cancer, in line with previous work (8,11,14,25). The one-year time frame was chosen to capture cancers that were present at the time of the blood being tested. Cancer stage was determined using the tumour, nodes, and metastasis (TNM) classification. Patients with a T stage of 3 or 4 and/or M stage of 1 were categorised as advanced stage.

### Exposure variables

Ethnicity was categorised into White, Asian, Black, Other, and Mixed in accordance with previous literature (20,21) and the 2021 UK census (26). A patient’s first blood test result in the study period was selected from the CPRD dataset; platelet count and (where recorded) CRP results were extracted from these records. Included platelet levels were within the range of 50 to 999×10^9^/L of blood and included CRP levels were between 0 to 500mg/L.

A high platelet count (thrombocytosis) was defined as a platelet count ≥400×10^9^/L blood in accordance with previous literature (8,11), and a high CRP level was defined as ≥7mg/L, calculated by determining the mean of the upper limit of normal from the laboratories in this study, as described previously (14).

### Statistical analysis

Multi-level logistic regression models, clustering patients within GP practices, were constructed to explore the association between a high test result to diagnosis of cancer within one year of test, stratified by test (platelet count or CRP). Differences in this association by ethnic group were explored by including an interaction term. The marginal distributions of the models were used to obtain adjusted cancer incidence estimates by group. Additional models assessed these effects in relation to the one-year incidence of cancers of specific sites, including colorectal, lung, oesophageal-gastric, and uterine, in accordance with the National Institute of Health and Care Excellence (NICE) guidelines (8).

Covariates included age group in 10-year bands (40 to 49 years, 50 to 59 years, 60 to 69 years, 70 to 79 years, and 80 years or above), sex (male or female, for cancers that are not sex-specific), smoking status (ever or never), tertile of the Cambridge Multimorbidity Score (CMS) ‘general outcome weighting’ (including an extra category for “no identified conditions”) (27), quintile of the area-based Index of Multiple Deprivation 2015 (IMD) (28), and body mass Index (BMI) (≤18.49 kg/m^2^, 18.5 to 24.99 kg/m^2^, 25.0 to 29.99 kg/m^2^, 30.0 to 39.99 kg/m^2^, ≥40 kg/m^2^). As different ethnic groups have higher incidences of certain cancer sites, the secondary analysis of stage at diagnosis also included an additional categorical covariate for cancer site.

### Sample size calculations

A sample size calculation determined that a sample size of 1,118 per group was sufficient to detect a cancer incidence of 3% with a margin of error of <1 percentage point.

### Patient and Public Involvement and Engagement

This study was developed in consultation with an existing Patient and Public Involvement and Engagement (PPIE) group, as part of a larger program of research. A dedicated group of three PPIE representatives were recruited for this study, comprising of both men and women, and ensuring representation from each of the main three ethnic groups analysed (White, Asian, and Black), and assisted in the interpretation of results.

### Reporting of results

Analyses were conducted using Stata MP version 18.0. Graphs were produced using Stata MP version 18.5 and GraphPad Prism version 10.2.3 for Windows. Results were reported in accordance with The Reporting of Studies Collected Using Observational Routinely-Collected Heath Data (RECORD) statement (29), as an extension of Strengthening and Reporting of Observational Studies in Epidemiology (STROBE) statement (30) (Supplementary materials 1).

## Results

### Patient selection process

Data were received from the CPRD for 5,796,384 patients who had a platelet count record, and 1,509,952 patients with a CRP record. After exclusions based on patient age, date of test, implausible blood test values or units of measure, there were 4,901,450 eligible patients with an acceptable platelet count record, and 811,559 eligible patients with an acceptable CRP record (Figure 1). Almost all the patients in the CRP cohort also had a recorded platelet count and so were also in the platelet cohort (804,086, 99%).

**Figure 1:**
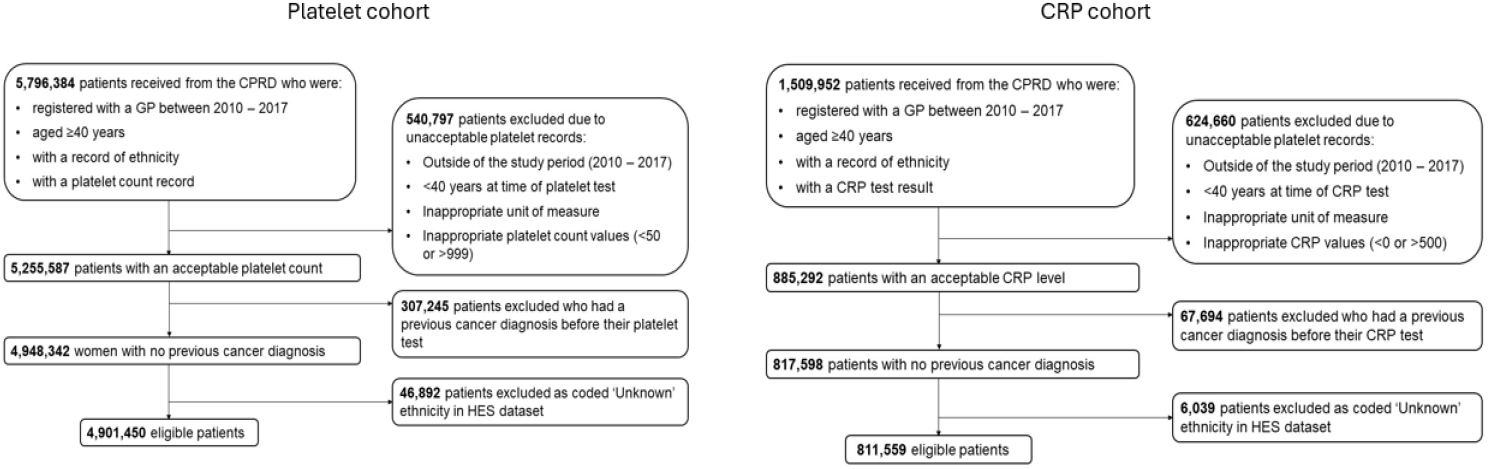
The cohort selection criteria to derive the platelet and CRP cohorts.

### Cohort characteristics

The distribution of patients across ethnic groups was broadly similar between the platelet and CRP cohort, with ≤1% of patients assigned to the Other or Mixed ethnic groups (Table 1). Results for patients from the Other and Mixed groups are detailed throughout the Supplementary materials.

**Table 1:** Cohort characteristic. *As measured by the Cambridge Multimorbidity Score. Overweight or obese BMI includes patients with a BMI of ≥25kg/m^2^.

| Ethnicity | n (%) | Sex,<br>% male | Age, median<br>(IQR) | Highest morbidity<br>burden<br>tertile*, % | Overweight<br>or obese<br>BMI, % | Most<br>deprived<br>quintile,<br>% |
| --- | --- | --- | --- | --- | --- | --- |
| **Platelet count** |  |  |  |  |  |  |
| White | 4,303,178<br>(87.8) | 45.3 | 58.8 (48.2-70.7) | 22.7 | 69.7 | 15.7 |
| Asian | 312,185 (6.4) | 47.4 | 50.3 (42.9-60.6) | 16.2 | 65.6 | 25.7 |
| Black | 192,444 (3.9) | 44.4 | 49.4 (43.7-58.4) | 12.8 | 79.0 | 44.0 |
| Other | 45,277 (<1) | 43.1 | 49.0 (43.3-57.7) | 12.0 | 70.6 | 28.2 |
| Mixed | 48,366 (<1) | 47.8 | 49.3 (43.2-59.0) | 10.8 | 71.2 | 28.3 |
| **CRP cohort** |  |  |  |  |  |  |
| White | 706,580<br>(87.1) | 41.8 | 61.0 (49.9-73.3) | 26.3 | 67.9 | 16.1 |
| Asian | 51,755 (6.4) | 42.4 | 54.3 (45.7-64.9) | 22.0 | 65.5 | 19.1 |
| Black | 37,140 (4.6) | 38.6 | 52.2 (46.0-63.0) | 18.3 | 78.5 | 43.3 |
| Other | 8,119 (1.0) | 38.8 | 51.0 (45.0-60.8) | 16.1 | 70.0 | 27.7 |
| Mixed | 7,965 (<1.0) | 43.1 | 51.3 (44.8-61.3) | 13.7 | 72.2 | 27.4 |

There was a slightly higher proportion of females in each ethnicity cohort (ranging from 52.2% for Mixed ethnicity to 56.9% for Other ethnicity). The White ethnic group were the oldest with a median age of 59. This compared to a median age of 49 or 50 years for non-White patients. Just under half of all Black patients in our cohort were assigned into the most deprived IMD quintile (44%), compared to only 15% of White patients. Black patients were also more likely to be overweight or obese (79%), compared to 66% to 72% of the remaining population.

### Distribution of platelet count in patients presenting to primary care by ethnicity

The proportion of patients with thrombocytosis generally increased with age (Figure 2a), with the exception of pre-menopausal women (aged 40 to 49 years) who had higher levels of thrombocytosis and higher median platelet levels than post-menopausal women, for all ethnic groups (Supplementary materials 2 and 3). For patients aged 50 years or above, the Black cohort had lower proportions of patients with thrombocytosis compared to White and Asian patients in the same age groups. A similar trend was seen across the ethnic groups for the distribution of median platelet count values (Supplementary material 3). A higher proportion of younger Asian patients (40 to 49 years) had thrombocytosis (4.5% (95% CI: 4.4 to 4.6%)) compared to younger patients from other ethnic groups, whereas a higher proportion of older White patients (80 years or above) had thrombocytosis (6.0% (95% CI 5.9 to 6.0%)). Women had higher proportions of thrombocytosis and higher median platelet count values compared to men, regardless of age and ethnicity (Supplementary materials 2 and 3)

**Figure 2:**
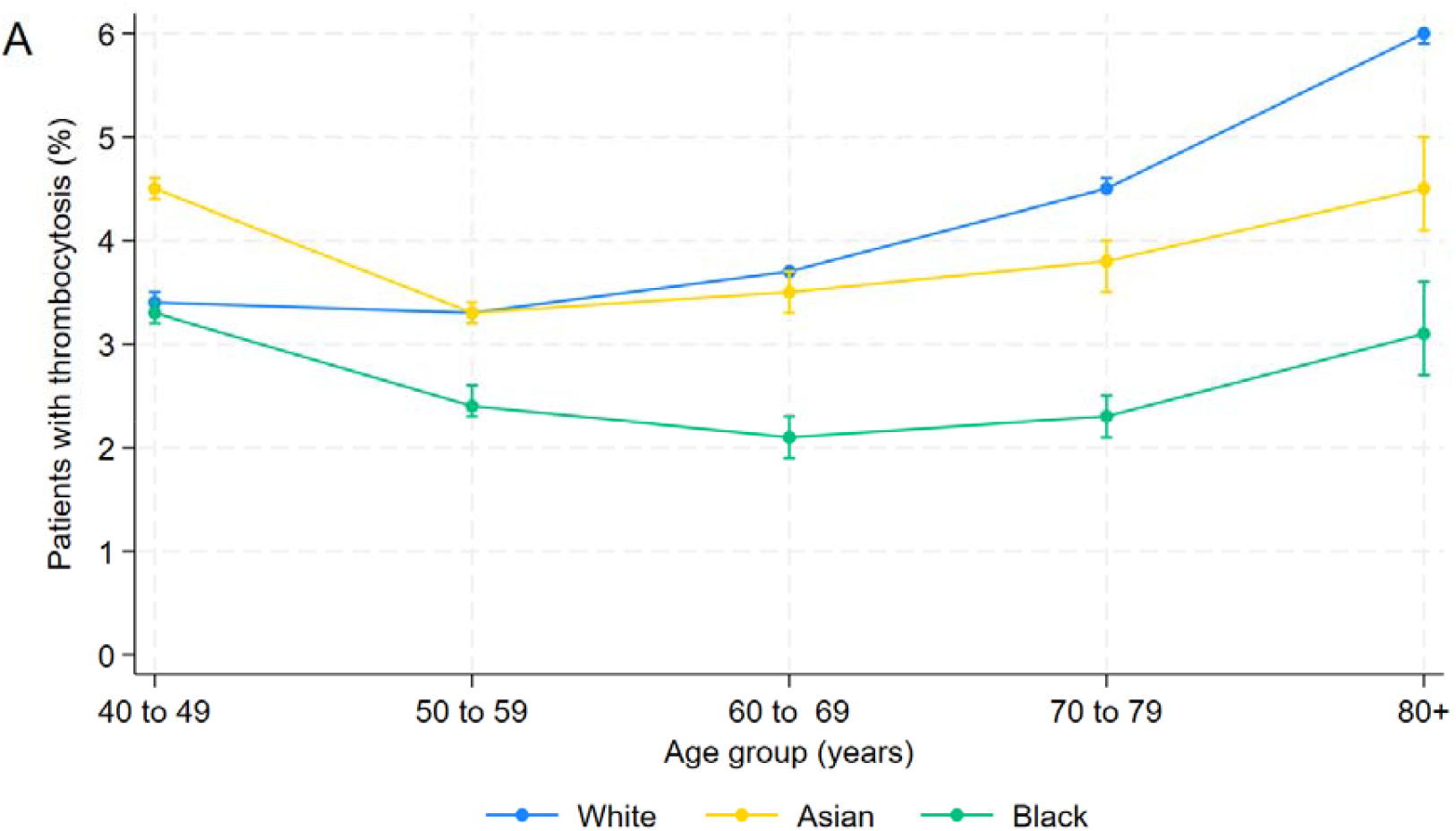
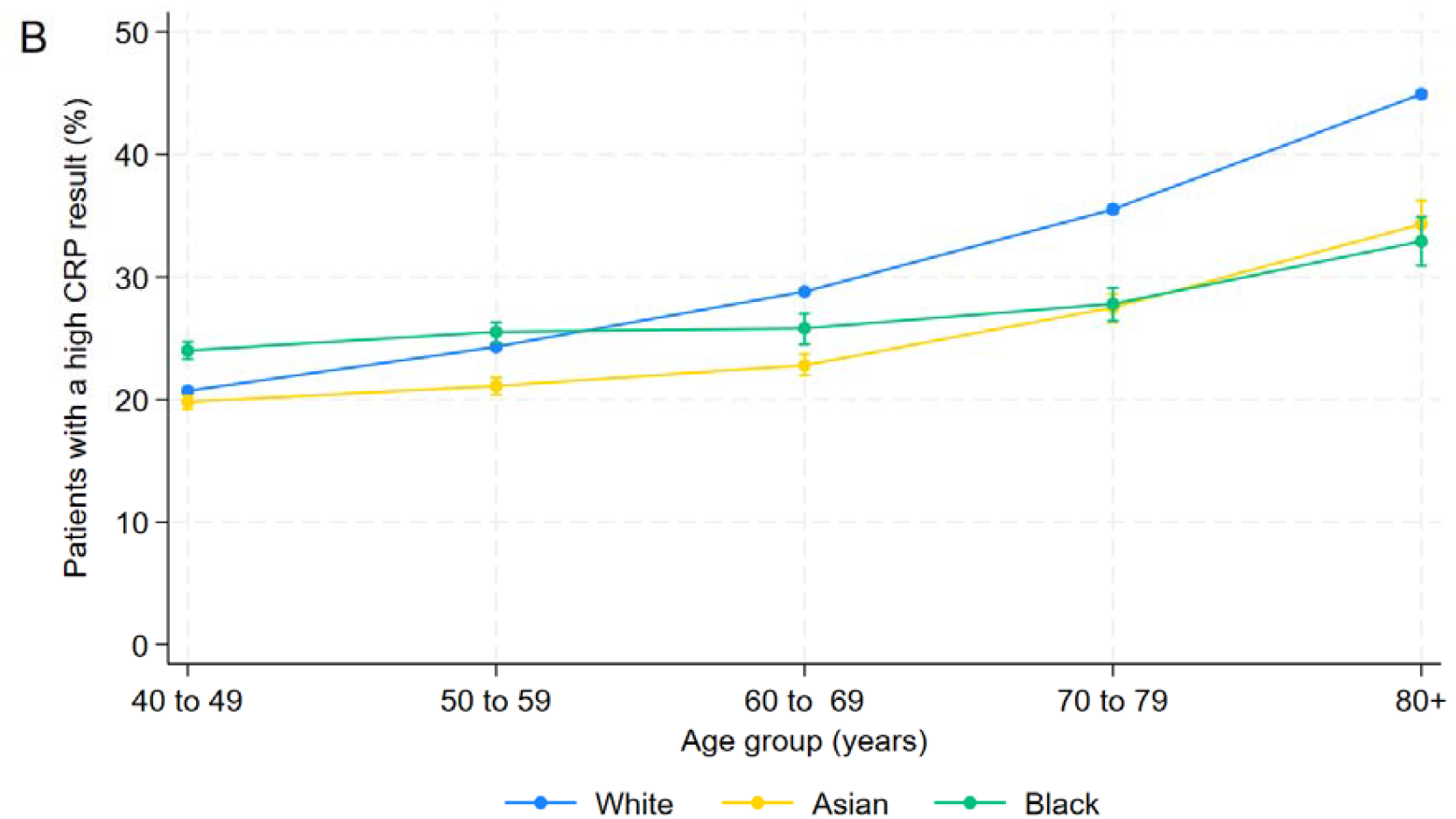
The percentage of patients with thrombocytosis (A), and a high CRP result (B), stratified by ethnicity and age group. Bars represent 95% CIs.

### Distribution of CRP level in patients presenting to primary care by ethnicity

The proportion of patients with a high CRP result increased with age, without a premenopausal spike for women as observed with thrombocytosis (Figure 2b, Supplementary materials 4). Black patients experienced a more gradual increase in with age than their White or Asian counterparts, which was reflected in a more constant distribution of median CRP values. White and Asian patients experienced a more pronounced rise with age (Supplementary materials 5). There was little evidence of sex differences of median CRP levels or proportion of patients with a high CRP result (Supplementary materials 4 and 5).

### One-year cancer incidence

Adjusted cancer incidence was highest for White patients, with the lowest rates found among Asian patients (Figure 3). The overall one-year cancer incidence for White and Black patients with thrombocytosis or a high CRP result exceeded 3%, though Asian patients had slightly lower incidence rates. There were no significant interactions between ethnicity, blood test result, and an incident cancer diagnosis, with similar diagnostic ORs observed across all ethnic groups, including Other and Mixed (Supplementary Materials 6 and 7). Unadjusted one-year cancer incidence rates are detailed in Supplementary Materials 8.

**Figure 3:**
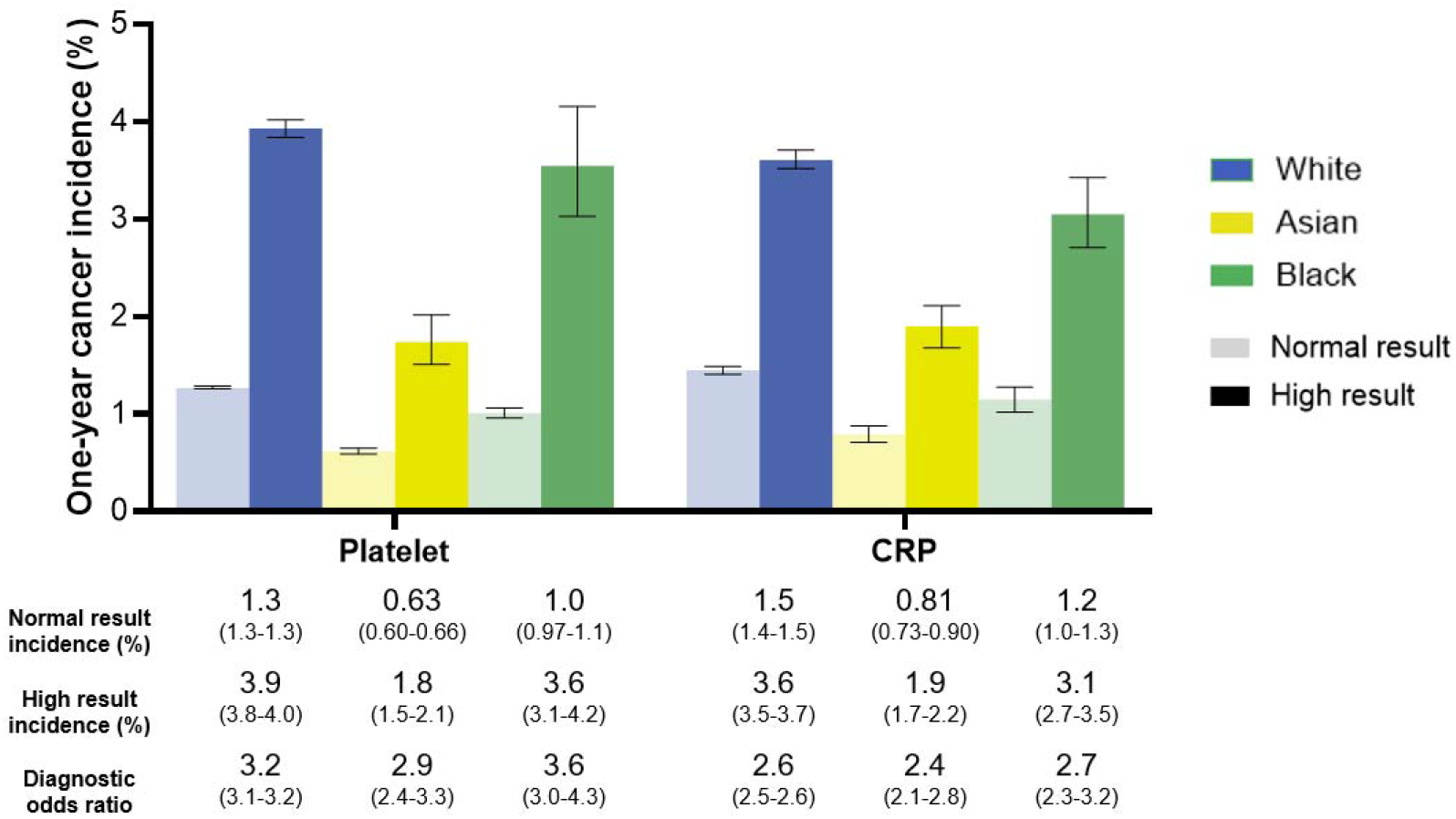
One-year cancer incidence by platelet count and CRP result, and ethnicity. Lighter bars represent a normal test result, and darker bars represent a high test result. Blue bars represent the average one-year cancer incidence for White patients, yellow bars for Asian patients, and green bars for Black patients. Black error bars represent 95% CIs. The table reports the incidence (%) and the diagnostic odds ratio of a one-year cancer diagnosis with 95% CIs in brackets underneath its corresponding ethnic group and blood marker from the chart above. Adjusted for gender, age group in 10-year bands (40 to 49 years, 50 to 59 years, 60 to 69 years, 70 to 79 years, and 80 + years), sex (male or female), smoking status (ever or never), CMS grouping, BMI category, IMD quintile, and year of test.

### One-year colorectal, lung, oesophageal-gastric, and uterine cancer incidence

Across colorectal, lung, oesophageal-gastric, and uterine cancers, thrombocytosis and a high CRP result were associated with higher one-year incidence compared with normal results (Figure 4). Exceptions were observed for Asian and Black women for uterine cancer by thrombocytosis status, and Asian patients for oesophageal-gastric cancer and Black women for uterine cancer by CRP status, where there was little evidence of increased cancer risk following a high test result.

**Figure 4:**
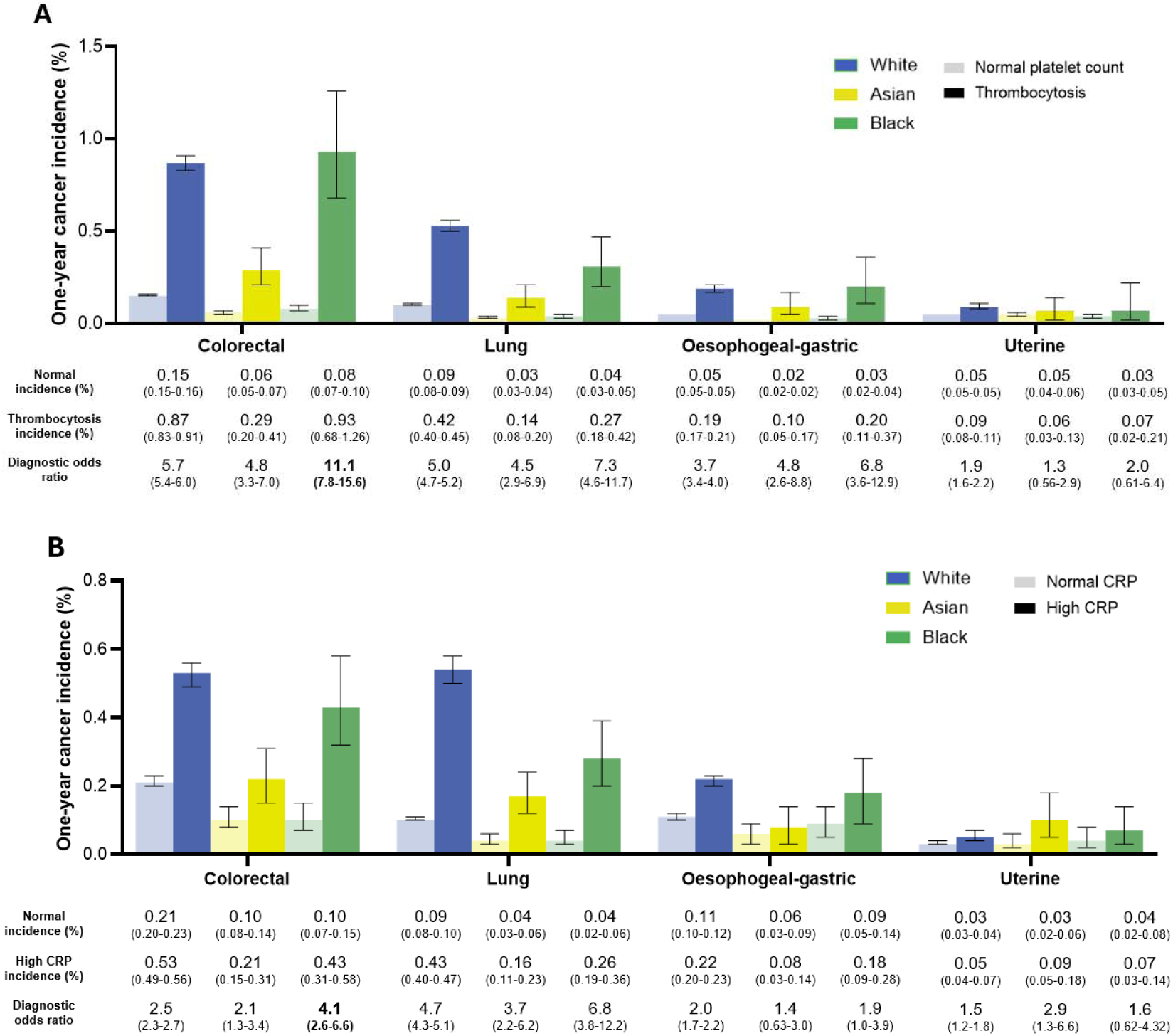
One-year colorectal, lung, oesophageal-gastric, and uterine cancer by platelet test result (A) / CRP result (B), and ethnicity. Lighter bars represent a normal test result, and darker bars represent a high result. Blue bars represent the average one-year cancer incidence for White patients, yellow bars for Asian patients, and green bars for Black patients. Black error bars represent 95% CIs. The table reports the incidence (%) and the diagnostic OR of a one-year cancer diagnosis with 95% CIs in brackets underneath its corresponding site and ethnic group from the chart above. The uterine cohorts included female patients only. Adjusted for gender, age group in 10-year bands (40 to 49 years, 50 to 59 years, 60 to 69 years, 70 to 79 years, and 80 + years), sex (male or female)*, smoking status (ever or never), CMS grouping, BMI category, IMD quintile, and year of test. * Sex was not included as a covariate for the ovarian cancer analyses.

Significant interaction effects were observed between ethnicity and blood test results for one-year colorectal cancer incidence (interaction *p*-value<0.001 for thrombocytosis and 0.043 for a high CRP result). Among Black patients, thrombocytosis was more associated with incident colorectal cancer (dOR 11.1, 95% CI 7.8 to 15.6%) compared to White patients (dOR 5.7, 95% CI 5.4 to 6.0, *p<*0.001), relative to a normal platelet count. A similar, although weaker, pattern was observed for raised CRP levels, with higher odds among Black patients (dOR 4.1, 95% CI 2.6 to 6.6) compared to White patients (dOR 2.5%, 95% CI 2.3 to 2.7%, *p*=0.043) (Figure 4).

Patients from Mixed ethnic groups with thrombocytosis, relative to normal platelet levels, had higher odds of incident oesophageal-gastric cancer compared to White patients (dOR 9.0, 95% CI 3.9 to 20.6 for Mixed; dOR 3.7, 3.4 to 4.0 for White; *p*=0.037). Similarly, women from Other groups with thrombocytosis had higher odds of incident uterine cancer compared to White women (dOR 13.7, 3.3 to 57.3 for Other; dOR 1.9, 95% CI 1.6 to 2.2; *p*=0.007). However, with only 8 Other women receiving a uterine cancer diagnosis, this result should be interpreted with caution (Supplementary Materials 6).

### One-year cancer incidence of other cancer sites

There was little evidence of ethnic differences in the interaction between ethnicity and blood test result and incident diagnoses of other cancer sites (Supplementary Materials 6 and 7). Notable exceptions included kidney cancer and leukaemia. For these cancers, thrombocytosis was more strongly associated with incident diagnosis, relative to a normal platelet count, among Black patients than among White patients (interaction p-values of 0.046 for kidney cancer and 0.004 for leukaemia). Some evidence of a similar pattern was observed among patients from Mixed ethnic groups for leukaemia, though this finding should be interpreted with caution given the small number of leukaemia diagnoses in this group (*n*=10) (Supplementary Materials 6).

### Advanced stage incidence

There was no evidence of differences in the proportion advanced stage cancers in the cohort of patients diagnosed with cancer in the year following their blood test by ethnic group (Figure 5). For both blood tests and for all ethnicities, a high result was more indicative of an advanced stage at diagnosis, however the evidence of this was weak for Asian patients with a high CRP result. There was no evidence of differences for the Other and Mixed groups (Supplementary materials 6 and 7).

**Figure 5:**
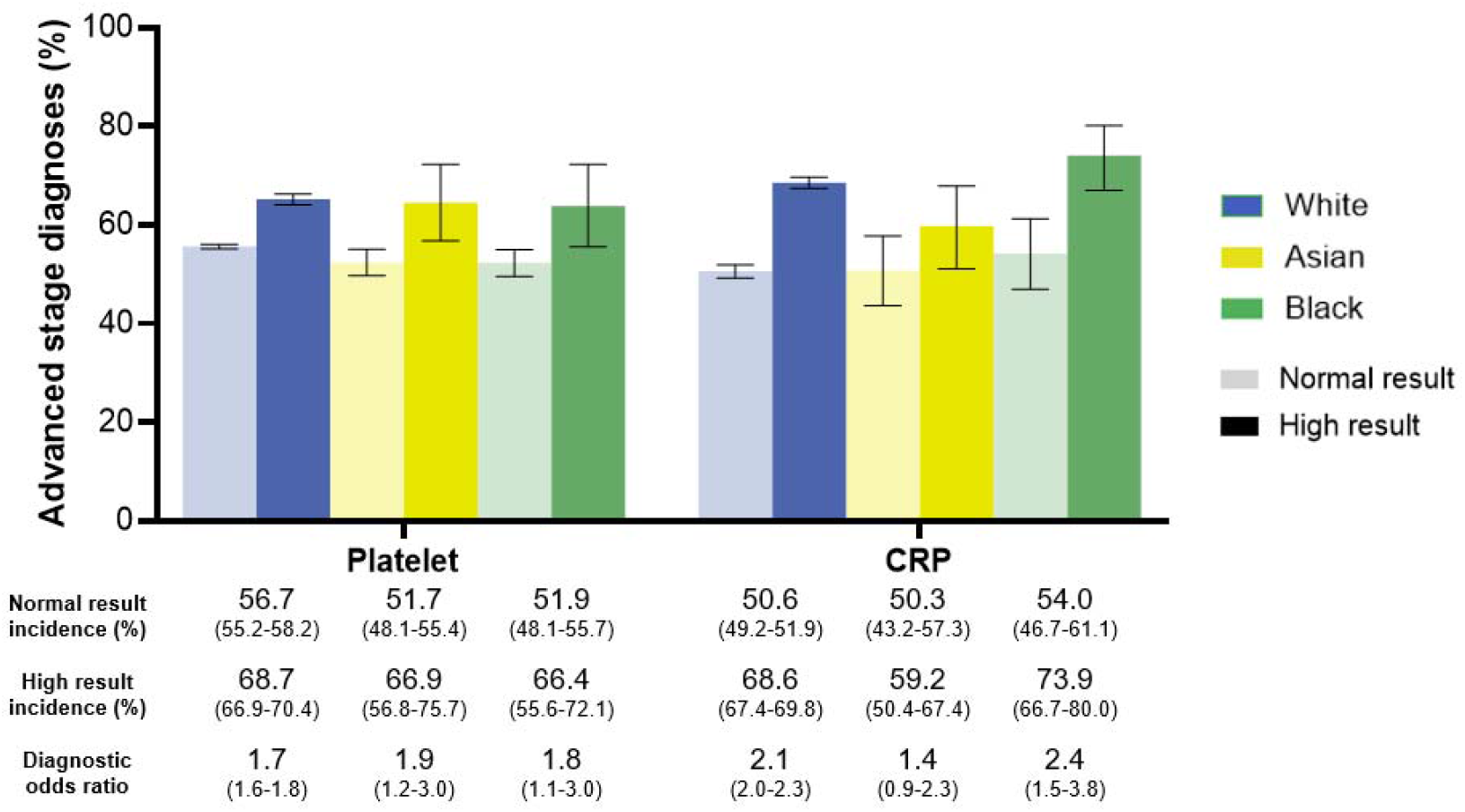
The proportion of cancer patients who were diagnosed at an advanced stage by platelet count and CRP result (normal or high) and ethnicity. Lighter bars represent a normal test result, and darker bars represent a high result. Blue bars represent the average proportion of White cancer patients diagnosed at an advanced stage, yellow bars represent the same for Asian patients, and green bars for Black patients. Black error bars represent 95% CIs. The table reports advanced stage diagnoses as a percentage (%) and the diagnostic OR of an advanced stage diagnosis for cancer patients is provided underneath its corresponding ethnic group and blood marker from the chart above, with 95% CIs. Adjusted for gender, age group in 10-year bands (40 to 49 years, 50 to 59 years, 60 to 69 years, 70 to 79 years, and 80 + years), sex (male or female), smoking status (ever or never), CMS grouping, BMI category, IMD quintile, and year of test.

## Discussion

This large UK primary care study found the predictive value for thrombocytosis and a raised CRP result for cancer incidence is generally consistent across ethnicities. There were site-specific differences: Black patients exhibited higher odds ratios of a colorectal, renal, and leukaemia diagnosis following a thrombocytosis result, relative to a normal platelet count, compared to White and Asian patients.

The overarching strength of this study was the unprecedented size of the cohort: just under five million patients who underwent a full blood count in primary care and over 800,000 with a CRP record. Such scale provides substantial statistical power but also carries the potential limitation of increased risk of incidental findings. There was little evidence of ethnic variation in the ability of thrombocytosis or a raised CRP result to detect one-year cancer incidence, which was largely consistent across each cancer site. There were three notable exceptions for thrombocytosis: colorectal, renal, and leukaemia. For these cancers, the dOR ratio associated with thrombocytosis, relative to a normal platelet count, was significantly higher for Black individuals compared with their White counterparts (interaction *p*-values <0.001, 0.046 and 0.004 for colorectal, kidney, and leukaemia, respectively). These findings must be interpreted cautiously, given the very large sample size and number of statistical comparisons performed, which increase the likelihood of chance associations. Nonetheless, the well-established links between thrombocytosis and both colorectal and renal cancer suggest that these differences could have genuine clinical relevance (9,11) In contrast, the limited evidence for an association between thrombocytosis and leukaemia warrants particular caution, and further work is needed to establish whether this represents a true biological signal, or a statistical artefact.

The potential for ethnicity-specific interpretation any biomarker raises important ethical considerations, particularly in relation to ethnicity-based adjustments to diagnostic thresholds (31). Such approaches can oversimplify the complex interplay of socio-economic, cultural, and structural determinants of health, and may inadvertently exacerbate health disparities rather than reduce them (31). Engagement with PPIE representatives highlighted that patient willingness and ability to engage with primary care services may differ substantially across ethnic groups, influenced by ethnicity-specific cultures and beliefs, religious practices, and socioeconomic status. Differences may also arise in whether and when blood tests are offered and accepted. These considerations are supported by a large body of qualitative research (32–40). A more responsible approach to evaluating diagnostic tools is to test and validate them in populations that are representative of those in which the tests are intended to be used. In the UK context, this means including ethnically diverse samples that reflect the nation’s multi-ethnic population, and any observed differences by ethnicity should be carefully investigated for their underlying causes, rather than assuming ethnicity alone is the determinant of risk.

The CPRD dataset delivered and analysed in this chapter is largely representative of the population of England and Wales in terms of ethnicity. According to the 2011 UK census (which is most relevant to the study period), 86.0% of the population were White, 7.5% were Asian, 3.3% were Black, 2.2% were Mixed, and 1% were from the Other ethnic group. The respective proportions represented in this chapter’s cohort were 87.8% White, 6.4% Asian, 3.9% Black, <1% Mixed, and <1% Other. The close alignment with national demographics, together with the large sample size, supports the generalisability of the findings revealed in this chapter. Importantly, it provides reassurance that the lack of observed ethnic differences in the predictive value of thrombocytosis for lung cancer supports its use as a universally applicable marker in primary care.

Whether these results are applicable to countries outside the UK is unclear. A recent systematic review found that Black individuals in the USA had higher platelet counts than their White counterparts (15), a direct contrast to the results found in this chapter. Different healthcare systems, access to healthcare, and cultures may contribute to these differences. Caution should be exercised in generalising the findings from this UK-based cohort to other international populations.

There are several limitations that should be considered. First, differences within ethnic groups, such as the differences between the Black African and Caribbean ethnic groups, or the East and South Asian ethnic groups, were not explored. The decision was made to align with common UK practices in ethnicity coding and to prioritise sample sizes, though the significance of heterogeneity within ethnic groups was recently highlighted by findings from the Covid-19 pandemic (41). Second, the use of area-based deprivation scores, such as the IMD, does not adequately account for variations of deprivations within areas, which have been shown to significantly differ within neighbourhoods by ethnicity (42). Third, it was not possible to determine the clinical rationale for ordering blood tests, whether patients from different ethnic groups present and report to primary care with equal frequency, or whether patients were offered and accepted blood tests equally. Fourth, emerging evidence from our research group suggests that using a patient’s first blood test result may underestimate cancer risk, and that using a random test would better reflect accurate cancer risk estimates. Nevertheless, this consideration does not affect the overall conclusions or clinical interpretation. Finally, much of NICE’s thrombocytosis guidance was defined in its 2015 update, meaning the majority of data in the present study were recorded before this guidance could have influenced clinical practice.

Overall, these findings underscore the need for ethnically diverse cohorts in evaluating diagnostic tests for cancer detection to avoid widening healthcare inequalities.

## Data Availability

The data that support the findings of this study are available from the CPRD (with linkage to HES and NCRAS). Restrictions apply to the availability of these data, which were used under license for the current study and so are not publicly available. Summary data are however available from the authors upon reasonable request and with permission of the CPRD, HES, and NCRAS.

## Authors’ contributions

LD and MB prepared the data for analysis. MB carried out the analysis and wrote the first draft of the manuscript. LM provided statistical support. SB TM LM and SWDM conceptualised the project and provided advice and supervision throughout. SWDM and JW provided clinical insights. All authors reviewed and revised the manuscript.

## Ethics approval

CPRD has ethics approval to support research using anonymised patient data from East Midlands - Derby Research Ethics Committee (ref 21/EM/0265). No individual patient data is included in this report.

## Acknowledgements

We thank our PPIE representatives Neomi Alam, Auguster Gold, and Andy Parsons. We acknowledge the work by Dr Sarah Price and Dr Bianca Wiering to provide code which enabled the calculation of patients’ CMS scores.

