## Supplementary materials 2 for "The impact of patient ethnicity on cancer incidence following platelet count and C-reactive protein tests in English primary care: a cohort study of 5 million patients"

Supplementary materials 2: Proportion of patients with thrombocytosis, stratified by ethnicity and age, reported as % (95% CI).

| **All** | **White** | **Asian** | **Black** | **Other** | **Mixed** |
| --- | --- | --- | --- | --- | --- |
| 40 to 49 years | 3.4 (3.4-3.5) | 4.5 (4.4-4.6) | 3.3 (3.2-3.4) | 3.7 (3.5-4.0) | 2.6 (2.4-2.8) |
| 50 to 59 years | 3.3 (3.3-3.3) | 3.3 (3.2-3.4) | 2.4 (2.3-2.6) | 3.0 (2.7-3.3) | 2.2 (1.9-2.4) |
| 60 to 69 years | 3.7 (3.7-3.7) | 3.5 (3.3-3.7) | 2.1 (1.9-2.3) | 2.9 (2.5-3.4) | 2.9 (2.5-3.3) |
| 70 to 79 years | 4.5 (4.5-4.6) | 3.8 (3.5-4.0) | 2.3 (2.1-2.5) | 2.8 (2.3-3.5) | 3.4 (2.8-4.1) |
| 80+ years | 6.0 (5.9-6.0) | 4.5 (4.1-5.0) | 3.1 (2.7-3.6) | 3.6 (2.7-4.7) | 4.7 (3.7-5.7) |

| **Males** | **White** | **Asian** | **Black** | **Other** | **Mixed** |
| --- | --- | --- | --- | --- | --- |
| 40 to 49 years | 2.0 (2.0-2.1) | 1.6 (1.5-1.7) | 1.2 (1.1-1.3) | 1.9 (1.6-2.2) | 1.2 (1.0-1.4) |
| 50 to 59 years | 2.3 (2.2-2.3) | 1.9 (1.7-2.0) | 1.7 (1.5-1.9) | 1.9 (1.5-2.3) | 1.4 (1.1-1.7) |
| 60 to 69 years | 2.6 (2.6-2.7) | 2.3 (2.1-2.5) | 1.8 (1.5-2.1) | 2.0 (1.5-2.7) | 1.8 (1.5-2.3) |
| 70 to 79 years | 2.9 (2.9-3.0) | 2.4 (2.1-2.6) | 1.9 (1.6-2.3) | 1.9 (1.2-2.7) | 2.2 (1.5-3.1) |
| 80+ years | 3.5 (3.4-3.6) | 2.8 (2.3-3.3) | 2.8 (2.2-3.5) | 1.5 (0.6-2.9) | 3.5 (2.2-5.1) |

| **Females** | **White** | **Asian** | **Black** | **Other** | **Mixed** |
| --- | --- | --- | --- | --- | --- |
| 40 to 49 years | 4.5 (4.4-4.5) | 7.0 (6.8-7.2) | 4.9 (4.7-5.0) | 5.0 (4.7-5.4) | 3.8 (3.5-4.1) |
| 50 to 59 years | 4.3 (4.3-4.4) | 4.7 (4.5-5.0) | 3.1 (2.9-3.3) | 4.0 (3.5-4.5) | 3.0 (2.5-3.5) |
| 60 to 69 years | 4.8 (4.7-4.8) | 4.5 (4.3-4.8) | 2.3 (2.0-2.6) | 3.6 (2.9-4.3) | 4.0 (3.3-4.7) |
| 70 to 79 years | 5.9 (5.8-6.0) | 5.1 (4.7-5.5) | 2.5 (2.2-2.9) | 3.6 (2.8-4.6) | 4.4 (3.4-5.5) |
| 80+ years | 7.3 (7.2-7.4) | 6.0 (5.3-6.6) | 3.4 (2.8-4.0) | 5.0 (3.6-6.7) | 5.4 (4.2-6.9) |
