## Supplementary materials 3 for "The impact of patient ethnicity on cancer incidence following platelet count and C-reactive protein tests in English primary care: a cohort study of 5 million patients"

Supplementary materials 3: Median platelet values by ethnicity and age group, reported as median (IQR), for all patients, males, and females.

| **All** | **White** | **Asian** | **Black** | **Other** | **Mixed** |
| --- | --- | --- | --- | --- | --- |
| 40 to 49 years | 254 (216-289) | 261 (221-308) | 238 (198-286) | 251 (212-298) | 248 (211-292) |
| 50 to 59 years | 252 (214-295) | 255 (216-299) | 234 (195-278) | 246 (208-289) | 241 (205-283) |
| 60 to 69 years | 247 (209-293) | 251(211-296) | 231 (191-276) | 241 (203-287) | 240 (202-282) |
| 70 to 79 years | 244 (204-293) | 245 (203-292) | 224 (185-269) | 230 (190-276) | 236 (197-282) |
| 80+ years | 246 (202-299) | 242 (199-293) | 219 (177-267) | 234 (193-280) | 235 (194-287) |

| **Males** | **White** | **Asian** | **Black** | **Other** | **Mixed** |
| --- | --- | --- | --- | --- | --- |
| 40 to 49 years | 240 (206-280) | 242 (207-281) | 219 (184-260) | 234 (198-274) | 234 (201-271) |
| 50 to 59 years | 238 (203-279) | 239 (204-280) | 220 (184-261) | 231 (194-269) | 229 (195-269) |
| 60 to 69 years | 232 (197-274) | 234 (198-275) | 214 (179-257) | 227 (191-268) | 224 (190-265) |
| 70 to 79 years | 224 (188-268) | 226 (190-270) | 208 (172-249) | 212 (177-251) | 220 (185-261) |
| 80+ years | 221 (183-269) | 223 (184-269) | 202 (164-247) | 215 (175-255) | 216 (175-261) |

| **Females** | **White** | **Asian** | **Black** | **Other** | **Mixed** |
| --- | --- | --- | --- | --- | --- |
| 40 to 49 years | 265 (226-310) | 280 (238-328) | 255 (212-304) | 265 (224-313) | 263 (224-309) |
| 50 to 59 years | 266 (228-309) | 271 (232-316) | 247 (208-292) | 260 (221-304) | 255 (218-298) |
| 60 to 69 years | 263 (225-308) | 267 (226-312) | 242 (202-286) | 254 (215-297) | 255 (218-299) |
| 70 to 79 years | 261 (221-309) | 264 (223-310) | 236 (198-280) | 248 (207-292) | 250 (211-298) |
| 80+ years | 259 (215-313) | 259 (214-309) | 233 (190-280) | 249 (207-298) | 246 (204-299) |
