## Supplementary materials 4 for "The impact of patient ethnicity on cancer incidence following platelet count and C-reactive protein tests in English primary care: a cohort study of 5 million patients"

Supplementary materials 4: Proportion of patients with a high CRP result, by ethnicity and age group, for all patients, males, and females. Reported as % (95% CI)

| **All** | **White** | **Asian** | **Black** | **Other** | **Mixed** |
| --- | --- | --- | --- | --- | --- |
| 40 to 49 years | 20.7 (20.5-20.8) | 19.8 (19.2-20.3) | 24.0 (23.3-24.7) | 19.4 (18.0-20.7) | 18.5 (17.2-19.8) |
| 50 to 59 years | 24.3 (24.1-24.5) | 21.1 (20.4-21.8) | 25.5 (24.6-26.3) | 22.8 (21.1-24.6) | 21.1 (19.4-22.9) |
| 60 to 69 years | 28.8 (28.6-29.0) | 22.8 (22.0-23.7) | 25.8 (24.5-27.0) | 25.3 (22.7-28.0) | 25.1 (22.6-27.6) |
| 70 to 79 years | 35.5 (35.2-35.8) | 27.5 (26.3-28.6) | 27.8 (26.4-29.1) | 29.0 (25.6-32.5) | 33.8 (30.1-37.6) |
| 80+ years | 44.9 (44.6-45.2) | 34.3 (32.6-36.2) | 32.9 (30.9-34.9) | 34.7 (30.0-40.0) | 41.3 (36.2-46.6) |

| **Male** | **White** | **Asian** | **Black** | **Other** | **Mixed** |
| --- | --- | --- | --- | --- | --- |
| 40 to 49 years | 21.9 (21.6-22.2) | 18.5 (17.6-19.3) | 20.6 (19.6-21.7) | 17.4 (15.5-19.4) | 17.8 (15.9-19.7) |
| 50 to 59 years | 24.7 (24.4-25.0) | 19.0 (18.0-20.1) | 22.3 (21.1-23.6) | 22.3 (19.6-25.2) | 21.4 (18.7-24.1) |
| 60 to 69 years | 30.0 (29.5-30.2) | 22.3 (21.0-23.6) | 25.4 (23.4-27.6) | 21.8 (18.0-26.1) | 24.1 (20.6-27.9) |
| 70 to 79 years | 37.8 (37.4-38.2) | 28.8 (27.1-30.5) | 29.8 (27.6-32.0) | 30.0 (24.9-35.4) | 34.5 (29.0-40.3) |
| 80+ years | 47.8 (47.3-48.3) | 34.5 (31.9-37.2) | 35.9 (32.8-39.1) | 33.4 (26.0-41.3) | 47.4 (38.8-56.2) |

| **Female** | **White** | **Asian** | **Black** | **Other** | **Mixed** |
| --- | --- | --- | --- | --- | --- |
| 40 to 49 years | 19.8 (19.5-20.0) | 20.9 (20.2-21.6) | 25.9 (25.0-26.8) | 20.2 (18.6-21.9) | 18.4 (16.8-20.2) |
| 50 to 59 years | 24.0 (23.8-24.3) | 22.6 (21.6-23.5) | 27.6 (26.5-28.7) | 23.2 (21.0-25.5) | 20.9 (18.7-23.3) |
| 60 to 69 years | 27.9 (27.6-28.2) | 23.2 (22.1-24.4) | 25.9 (24.4-27.5) | 27.5 (24.1-31.1) | 25.9 (18.7-23.3) |
| 70 to 79 years | 33.7 (33.4-34.0) | 26.3 (24.8-27.9) | 26.4 (24.7-28.1) | 28.2 (23.7-33.0) | 33.2 (28.3-38.5) |
| 80+ years | 43.3 (42.9-43.7) | 34.3 (31.9-36.7) | 30.6 (28.1-33.3) | 35.6 (29.3-42.3) | 27.8 (31.4-44.4) |
