## Supplementary materials 5 for "The impact of patient ethnicity on cancer incidence following platelet count and C-reactive protein tests in English primary care: a cohort study of 5 million patients"

Supplementary materials 5: Median CRP values by ethnicity and age group for all patients, males, and females, reported as median (IQR).

| **All** | **White** | **Asian** | **Black** | **Other** | **Mixed** |
| --- | --- | --- | --- | --- | --- |
| 40 to 49 years | 3.2 (1.5-5.0) | 3.3 (1.3-5.0) | 5.0 (2.0-6.1) | 4.0 (1.9-5.0) | 3.6 (1.6-5.0) |
| 50 to 59 years | 4.0 (2.0-6.3) | 4.0 (1.7-5.7) | 5.0 (2.0-7.0) | 4.0 (2.0-6.0) | 4.0 (2.0-5.5) |
| 60 to 69 years | 4.0 (2.0-8.0) | 4.0 (1.7-6) | 5.0 (2.0-7.0) | 4.0 (2.0-7.0) | 4.0 (2.0-7.0) |
| 70 to 79 years | 4.6 (2.0-11.4) | 4.0 (1.7-7.6) | 5.0 (2.0-8.0) | 4.0 (2.0-8.0) | 5.0 (2.0-10.0) |
| 80+ years | 5.0 (3.0-19.0) | 5.0 (2.0-11.0) | 5.0 (2.0-10.2) | 5.0 (2.0-11.0) | 5.0 (3.0-17.0) |

| **Males** | **White** | **Asian** | **Black** | **Other** | **Mixed** |
| --- | --- | --- | --- | --- | --- |
| 40 to 49 years | 3.7 (1.9-5.7) | 3.0 (1.2-5.0) | 4.0 (2.0-5.0) | 3.7 (1.8-5.0) | 3.1 (1.6-5.0) |
| 50 to 59 years | 4.0 (2.0-6.6) | 3.0 (1.0-5.0) | 4.0 (2.0-6.0) | 3.6 (2.0-5.8) | 3.4 (2.0-5.0) |
| 60 to 69 years | 4.0 (2.0-9.0) | 3.4 (1.0-5.6) | 5.0 (2.0-7.0) | 4.0 (2.0-6.0) | 4.0 (2.0-6.4) |
| 70 to 79 years | 5.0 (2.0-14.0) | 4.0 (1.3-9.0) | 5.0 (2.0-8.9) | 4.0 (2.0-9.0) | 5.0 (2.0-11.0) |
| 80+ years | 6.0 (3.0-23.0) | 5.0 (2.0-12.0) | 5.0 (2.0-12.7) | 5.0 (2.0-10.5) | 5.7 (3.0-20.0) |

| **Females** | **White** | **Asian** | **Black** | **Other** | **Mixed** |
| --- | --- | --- | --- | --- | --- |
| 40 to 49 years | 3.0 (1.2-5.0) | 4.0 (1.5-5.6) | 5.0 (2.0-7.0) | 4.0 (1.8-5.0) | 3.9 (1.7-5.0) |
| 50 to 59 years | 4.0 (2.0-6.1) | 4.0 (2.0-6.0) | 5.0 (2.0-7.1) | 4.0 (2.0-6.0) | 4.0 (2.0-6.0) |
| 60 to 69 years | 4.0 (2.0-7.9) | 4.0 (2.0-6.0) | 5.0 (2.0-7.0) | 4.0 (2.0-7.8) | 4.3 (2.0-7.0) |
| 70 to 79 years | 4.1 (2.0-10.0) | 4.0 (2.0-7.0) | 5.0 (2.0-7.0) | 4.1 (2.0-8.0) | 5.0 (2.8-9.2) |
| 80+ years | 5.0 (2.9-17.0) | 4.9 (2.0-10.7) | 5.0 (2.0-9.0) | 5.0 (2.0-11.1) | 5.0 (3.0-12.0) |
