## Supplementary materials 6 for "The impact of patient ethnicity on cancer incidence following platelet count and C-reactive protein tests in English primary care: a cohort study of 5 million patients"

**Supplementary 6.** **One-year cancer incidence by platelet test result and ethnicity.**

Analyses were adjusted for sex, age group in 10-year bands (40 to 49 years, 50 to 59 years, 60 to 69 years, 70 to 79 years, and 80 + years), smoking status, CMS grouping, BMI category, IMD quintile, and year of test.

| **All cancer** | | | | | |
| --- | --- | --- | --- | --- | --- |
|  | *n* | Normal test result incidence, % (95% CI) | High test result incidence, % (95% CI) | Interaction p-value | Odds ratio (95% CI) |
| White | 85,552 | 1.27 (1.25 to 1.29) | 3.91 (3.82 to 4.00) | - | 3.16 (3.09 to 3.23) |
| Asian | 2,093 | 0.63 (0.60 to 0.66) | 1.78 (1.54 to 2.05) | 0.184 | 2.85 (2.44 to 3.32) |
| Black | 1,950 | 1.02 (0.97 to 1.07) | 3.58 (3.06 to 4.19) | 0.145 | 3.59 (3.03 to 4.26) |
| Other | 462 | 1.02 (0.92 to 1.12) | 3.71 (2.77 to 4.95) | 0.286 | 3.76 (2.74 to 5.15) |
| Mixed | 495 | 0.94 (0.82 to 1.03) | 3.87 (2.93 to 5.10) | 0.053 | 4.27 (3.15 to 5.78) |
| **Advanced stage** | | | | | |
|  | *n* | Normal test result incidence, % (95% CI) | High test result incidence, % (95% CI) | Interaction p-value | Odds ratio (95% CI) |
| White | 29,931 | 56.7 (55.2 to 58.2) | 68.7 (66.9 to 70.4) | - | 1.68 (1.57 to 1.79) |
| Asian | 596 | 51.7 (48.1 to 55.4) | 66.9 (56.8 to 75.7) | 0.589 | 1.89 (1.20 to 2.96) |
| Black | 558 | 51.9 (48.1 to 55.7) | 66.4 (55.6 to 75.8) | 0.694 | 1.84 (1.14 to 2.95) |
| Other | 151 | 52.9 (45.8 to 59.8) | 76.3 (57.7 to 88.4) | 0.231 | 2.88 (1.17 to 7.10) |
| Mixed | 193 | 57.7 (51.1 to 64.0) | 83.3 (66.0 to 92.8) | 0.123 | 3.66 (1.38 to 9.72) |
| **Colorectal** | | | | | |
|  | *n* | Normal test result incidence, % (95% CI) | High test result incidence, % (95% CI) | Interaction p-value | Odds ratio (95% CI) |
| White | 12,435 | 0.15 (0.15 to 0.16) | 0.87 (0.83 to 0.91) | - | 5.69 (5.43 to 5.96) |
| Asian | 229 | 0.06 (0.05 to 0.07) | 0.29 (0.20 to 0.41) | 0.370 | 4.79 (3.29 to 6.96) |
| Black | 197 | 0.08 (0.07 to 0.10) | 0.93 (0.68 to 1.26) | **>0.001** | **11.1 (7.82 to 15.6)** |
| Other | 47 | 0.09 (0.06 to 0.12) | 0.72 (0.37 to 1.37) | 0.334 | 8.16 (3.93 to 17.0) |
| Mixed | 64 | 0.10 (0.08 to 0.13) | 0.85 (0.48 to 1.49) | 0.243 | 8.31 (4.41 to 15.7) |
| **Lung** | | | | | |
|  | *n* | Normal test result incidence, % (95% CI) | High test result incidence, % (95% CI) | Interaction p-value | Odds ratio (95% CI) |
| White | 10,955 | 0.09 (0.08 to 0.09) | 0.42 (0.40 to 0.45) | - | 4.98 (4.74 to 5.23) |
| Asian | 167 | 0.03 (0.03 to 0.04) | 0.14 (0.09 to 0.20) | 0.630 | 4.46 (2.87 to 6.94) |
| Black | 131 | 0.04 (0.03 to 0.05) | 0.27 (0.18 to 0.42) | 0.110 | 7.32 (4.57 to 11.7) |
| Other | 47 | 0.06 (0.04 to 0.08) | 0.42 (0.21 to 0.84) | 0.411 | 6.87 (3.19 to 14.8) |
| Mixed | 45 | 0.05 (0.04 to 0.07) | 0.33 (0.16 to 0.69) | 0.605 | 6.17 (2.73 to 14.0) |
| **Oesophageal-gastric** | | | | | |
|  | *n* | Normal test result incidence, % (95% CI) | High test result incidence, % (95% CI) | Interaction p-value | Odds ratio (95% CI) |
| White | 4,530 | 0.05 (0.05 to 0.05) | 0.19 (0.17 to 0.21) | - | 3.67 (3.36 to 4.01) |
| Asian | 91 | 0.02 (0.02 to 0.02) | 0.10 (0.05 to 0.17) | 0.390 | 4.81 (2.62 to 8.83) |
| Black | 80 | 0.03 (0.02 to 0.04) | 0.20 (0.11 to 0.37) | 0.062 | 6.79 (3.58 to 12.9) |
| Other | 22 | 0.04 (0.02 to 0.06) | 0.29 (0.11 to 0.79) | 0.146 | 8.24 (2.78 to 24.4) |
| Mixed | 36 | 0.05 (0.03 to 0.07) | 0.44 (0.21 to 0.93) | **0.037** | **8.96 (3.90 to 20.6)** |
| **Uterine** | | | | | |
|  | *n* | Normal test result incidence, % (95% CI) | High test result incidence, % (95% CI) | Interaction p-value | Odds ratio (95% CI) |
| White | 1,826 | 0.05 (0.05 to 0.05) | 0.09 (0.08 to 0.11) | - | 1.87 (1.60 to 2.20) |
| Asian | 89 | 0.05 (0.04 to 0.06) | 0.06 (0.03 to 0.13) | 0.373 | 1.28 (0.56 to 2.93) |
| Black | 50 | 0.03 (0.03 to 0.05) | 0.07 (0.02 to 0.21) | 0.931 | 1.97 (0.61 to 6.35) |
| Other | 8 | 0.02 (0.01 to 0.04) | 0.24 (0.08 to 0.74) | **0.007** | **13.7 (3.25 to 57.3)** |
| Mixed | 9 | 0.03 (0.01 to 0.05) | 0.08 (0.01 to 0.60) | 0.631 | 3.13 (0.39 to 25.1) |
| **Prostate** | | | | | |
|  | *n* | Normal test result incidence, % (95% CI) | High test result incidence, % (95% CI) | Interaction p-value | Odds ratio (95% CI) |
| White | 13,558 | 0.29 (0.28 to 0.30) | 0.37 (0.33 to 0.40) | - | 1.26 (1.15 to 1.38) |
| Asian | 222 | 0.11 (0.09 to 0.12) | 0.16 (0.07 to 0.33) | 0.678 | 1.48 (0.69 to 3.14) |
| Black | 619 | 0.58 (0.53 to 0.64) | 0.91 (0.57 to 1.45) | 0.367 | 1.57 (0.97 to 2.53) |
| Other | 95 | 0.38 (0.31 to 0.47) | 0.62 (0.20 to 1.91) | 0.661 | 1.63 (0.51 to 5.22) |
| Mixed | 82 | 0.25 (0.20 to 0.31) | 0.30 (0.07 to 1.20) | 0.940 | 1.19 (0.29 to 4.89) |
| **Breast** | | | | | |
|  | *n* | Normal test result incidence, % (95% CI) | High test result incidence, % (95% CI) | Interaction p-value | Odds ratio (95% CI) |
| White | 7,812 | 0.30 (0.29 to 0.31) | 0.31 (0.29 to 0.35) | - | 1.05 (0.95 to 1.16) |
| Asian | 329 | 0.21 (0.19 to 0.24) | 0.26 (0.17 to 0.40) | 0.465 | 1.24 (0.80 to 1.91) |
| Black | 188 | 0.19 (0.17 to 0.22) | 0.36 (0.20 to 0.63) | 0.057 | 1.87 (1.04 to 3.35) |
| Other | 59 | 0.27 (0.21 to 0.34) | 0.10 (0.01 to 0.72) | 0.320 | 0.38 (0.05 to 2.78) |
| Mixed | 51 | 0.21 (0.16 to 0.28) | 0.34 (0.11 to 1.06) | 0.465 | 1.62 (0.50 to 5.23) |
| **Lymphoma** | | | | | |
|  | *n* | Normal test result incidence, % (95% CI) | High test result incidence, % (95% CI) | Interaction p-value | Odds ratio (95% CI) |
| White | 4,132 | 0.07 (0.07 to 0.07) | 0.17 (0.16 to 0.19) | - | 2.48 (2.22 to 2.76) |
| Asian | 154 | 0.05 (0.04 to 0.06) | 0.10 (0.05 to 0.18) | 0.552 | 2.03 (1.07 to 3.86) |
| Black | 95 | 0.05 (0.04 to 0.07) | 0.15 (0.07 to 0.33) | 0.815 | 2.73 (1.20 to 6.25) |
| Other | 23 | 0.05 (0.03 to 0.08) | 0.24 (0.08 to 0.76) | 0.260 | 4.99 (1.48 to 16.8) |
| Mixed | 38 | 0.08 (0.06 to 0.11) | 0.16 (0.04 to 0.63) | 0.808 | 2.07 (0.50 to 8.63) |
| **Pancreas** | | | | | |
|  | *n* | Normal test result incidence, % (95% CI) | High test result incidence, % (95% CI) | Interaction p-value | Odds ratio (95% CI) |
| White | 3,123 | 0.07 (0.06 to 0.07) | 0.11 (0.09 to 0.12) | - | 1.60 (1.39 to 1.84) |
| Asian | 53 | 0.02 (0.02 to 0.03) | 0.05 (0.00 to 0.10) | 0.523 | 2.24 (0.81 to 6.21) |
| Black | 77 | 0.07 (0.05 to 0.08) | . | . | . |
| Other | 19 | 0.07 (0.04 to 0.10) | 0.12 (-0.11 to 0.35) | 0.919 | 1.78 (0.24 to 13.4) |
| Mixed | 17 | 0.05 (0.03 to 0.08) | 0.10 (-0.10 to 0.30) | 0.821 | 2.02 (0.27 to 15.3) |
| **Leukaemia** | | | | | |
|  | *n* | Normal test result incidence, % (95% CI) | High test result incidence, % (95% CI) | Interaction p-value | Odds ratio (95% CI) |
| White | 2,711 | 0.06 (0.06 to 0.06) | 0.08 (0.07 to 0.10) | - | 1.41 (1.20 to 1.67) |
| Asian | 79 | 0.03 (0.02 to 0.04) | 0.09 (0.02 to 0.16) | 0.073 | 2.92 (1.34 to 6.35) |
| Black | 41 | 0.03 (0.02 to 0.04) | 0.16 (0.02 to 0.30) | **0.004** | **5.78 (2.27 to 14.8)** |
| Other | 17 | 0.06 (0.03 to 0.08) | . | . | 1.41 (1.20 to 1.67) |
| Mixed | 10 | 0.02 (0.01 to 0.04) | 0.21 (0 to 0.50) | **0.017** | **9.40 (1.99 to 44.4)** |
| **Bladder** | | | | | |
|  | *n* | Normal test result incidence, % (95% CI) | High test result incidence, % (95% CI) | Interaction p-value | Odds ratio (95% CI) |
| White | 2,749 | 0.06 (0.05 to 0.06) | 0.15 (0.13 to 0.17) | - | 2.62 (2.30 to 2.98) |
| Asian | 41 | 0.02 (0.01 to 0.03) | 0.02 (-0.02 to 0.05) | 0.264 | 0.84 (0.12 to 6.12) |
| Black | 14 | 0.01 (0.01 to 0.02) | 0.04 (-0.04 to 0.12) | 0.815 | 3.34 (0.44 to 25.5) |
| Other | 4 | 0.02 (0.00 to 0.03) | . | . | . |
| Mixed | 8 | 0.03 (0.01 to 0.05) | . | . | . |
| **Kidney** | | | | | |
|  | *n* | Normal test result incidence, % (95% CI) | High test result incidence, % (95% CI) | Interaction p-value | Odds ratio (95% CI) |
| White | 2,611 | 0.04 (0.04 to 0.04) | 0.18 (0.16 to 0.20) | - | 4.70 (4.21 to 5.2) |
| Asian | 67 | 0.02 (0.01 to 0.02) | 0.11 (0.06 to 0.20) | 0.415 | 6.18 (3.23 to 11.8) |
| Black | 41 | 0.02 (0.01 to 0.02) | 0.18 (0.09 to 0.37) | **0.046** | 10.4 (4.80 to 22.6) |
| Other | 15 | 0.03 (0.02 to 0.05) | 0.08 (0.01 to 0.58) | 0.556 | 2.55 (0.33 to 19.4) |
| Mixed | 22 | 0.04 (0.02 to 0.06) | 0.25 (0.08 to 0.76) | 0.600 | 6.52 (1.93 to 22.1) |
| **Ovarian** | | | | | |
|  | *n* | Normal test result incidence, % (95% CI) | High test result incidence, % (95% CI) | Interaction p-value | Odds ratio (95% CI) |
| White | 2,144 | 0.06 (0.06 to 0.06) | 0.34 (0.31 to 0.38) | - | 5.71 (5.16 to 6.32) |
| Asian | 63 | 0.03 (0.02 to 0.04) | 0.15 (0.09 to 0.26) | 0.777 | 5.23 (2.89 to 9.49) |
| Black | 39 | 0.03 (0.02 to 0.05) | 0.18 (0.08 to 0.40) | 0.861 | 5.28 (2.21 to 12.6) |
| Other | 14 | 0.04 (0.02 to 0.07) | 0.48 (0.20 to 1.16) | 0.136 | 13.18 (4.41 to 39.4) |
| Mixed | 8 | 0.02 (0.01 to 0.05) | 0.20 (0.05 to 0.80) | 0.604 | 8.73 (1.76 to 43.3) |
| **Myeloma** | | | | | |
|  | *n* | Normal test result incidence, % (95% CI) | High test result incidence, % (95% CI) | Interaction p-value | Odds ratio (95% CI) |
| White | 1,721 | 0.04 (0.04 to 0.04) | 0.04 (0.03 to 0.05) | - | 1.17 (0.94 to 1.46) |
| Asian | 73 | 0.03 (0.02 to 0.04) | 0.04 (0 to 0.09) | 0.928 | 1.24 (0.39 to 3.93) |
| Black | 99 | 0.08 (0.06 to 0.10) | 0.10 (0 to 0.22) | 0.872 | 1.29 (0.41 to 4.08) |
| Other | 12 | 0.04 (0.02 to 0.07) | . | . | . |
| Mixed | 8 | 0.02 (0.01 to 0.04) | 0.10 (0 to 0.30) | 0.181 | 4.94 (0.61 to 40.2) |
| **Oral** | | | | | |
|  | *n* | Normal test result incidence, % (95% CI) | High test result incidence, % (95% CI) | Interaction p-value | Odds ratio (95% CI) |
| White | 1,798 | 0.04 (0.04 to 0.04) | 0.06 (0.05 to 0.07) | - | 1.43 (1.17 to 1.76) |
| Asian | 71 | 0.02 (0.02 to 0.03) | 0.03 (0.00 to 0.07) | 0.966 | 1.40 (0.44 to 4.44) |
| Black | 25 | 0.01 (0.01 to 0.02) | 0.05 (0 to 0.11) | 0.236 | 3.46 (0.81 to 14.7) |
| Other | 9 | 0.02 (0.01 to 0.04) | . | . | . |
| Mixed | 9 | 0.02 (0.01 to 0.03) | 0.09 (0 to 0.27) | 0.223 | 5.25 (0.66 to 42.1) |
| **Head and Neck** | | | | | |
|  | *n* | Normal test result incidence, % (95% CI) | High test result incidence, % (95% CI) | Interaction p-value | Odds ratio (95% CI) |
| White | 1,205 | 0.03 (0.03 to 0.03) | 0.04 (0.03 to 0.05) | - | 1.47 (1.16 to 1.88) |
| Asian | 79 | 0.03 (0.02 to 0.03) | 0.04 (0.01 to 0.08) | 0.737 | 1.73 (0.70 to 4.28) |
| Black | 38 | 0.02 (0.01 to 0.03) | . | . | . |
| Other | 11 | 0.03 (0.01 to 0.04) | . | . | . |
| Mixed | 12 | 0.02 (0.01 to 0.04) | 0.08 (-0.08 to 0.23) | 0.428 | 3.39 (0.44 to 26.3) |
