## Supplementary materials 7 for "The impact of patient ethnicity on cancer incidence following platelet count and C-reactive protein tests in English primary care: a cohort study of 5 million patients"

**Supplementary materials 7: Cancer incidence and odds ratios (CRP)**

| **All cancer** | | | | | |
| --- | --- | --- | --- | --- | --- |
|  | *n* | Normal test result incidence, % (95% CI) | High test result incidence, % (95% CI) | Interaction p-value | Odds ratio (95% CI) |
| White | 20,650 | 1.45 (1.41 to 1.49) | 3.59 (3.50 to 3.69) | - | 2.54 (2.47 to 2.61) |
| Asian | 607 | 0.81 (0.73 to 0.90) | 1.93 (1.70 to 2.19) | 0.542 | 2.41 (2.05 to 2.84) |
| Black | 603 | 1.16 (1.04 to 1.30) | 3.08 (2.74 to 3.47) | 0.439 | 2.71 (2.30 to 3.19) |
| Other | 124 | 1.16 (0.92 to 1.47) | 2.93 (2.24 to 3.82) | 0.944 | 2.57 (1.79 to 3.70) |
| Mixed | 105 | 0.90 (0.69 to 1.18) | 2.61 (1.99 to 3.42) | 0.445 | 2.95 (2.00 to 4.35) |
| **Advanced stage** | | | | | |
|  | *n* | Normal test result incidence, % (95% CI) | High test result incidence, % (95% CI) | Interaction p-value | Odds ratio (95% CI) |
| White | 8,352 | 50.55 (49.20 to 51.91) | 68.62 (67.42 to 69.79) | - | 2.14 (1.98 to 2.31) |
| Asian | 202 | 50.26 (43.18 to 57.32) | 59.16 (50.42 to 67.35) | 0.087 | 1.43 (0.91 to 2.25) |
| Black | 243 | 54.00 (46.72 to 61.12) | 73.89 (66.68 to 80.00) | 0.605 | 2.41 (1.54 to 3.77) |
| Other | 42 | 39.69 (26.05 to 55.14) | 66.84 (48.91 to 80.92) | 0.470 | 3.06 (1.16 to 8.09) |
| Mixed | 36 | 53.77 (36.28 to 70.37) | 49.09 (31.03 to 67.39) | 0.076 | 0.83 (0.29 to 2.36) |
| **Colorectal** | | | | | |
|  | *n* | Normal test result incidence, % (95% CI) | High test result incidence, % (95% CI) | Interaction p-value | Odds ratio (95% CI) |
| White | 3,211 | 0.21 (0.20 to 0.23) | 0.53 (0.49 to 0.56) | - | 2.52 (2.34 to 2.70) |
| Asian | 76 | 0.10 (0.08 to 0.14) | 0.21 (0.15 to 0.31) | 0.474 | 2.12 (1.34 to 3.36) |
| Black | 70 | 0.10 (0.07 to 0.15) | 0.43 (0.31 to 0.58) | **0.043** | 4.10 (2.57 to 6.57) |
| Other | 11 | 0.06 (0.02 to 0.17) | 0.36 (0.17 to 0.76) | 0.199 | 5.64 (1.65 to 19.3) |
| Mixed | 10 | 0.11 (0.05 to 0.24) | 0.14 (0.05 to 0.44) | 0.305 | 1.24 (0.32 to 4.79) |
| **Lung** | | | | | |
|  | *n* | Normal test result incidence, % (95% CI) | High test result incidence, % (95% CI) | Interaction p-value | Odds ratio (95% CI) |
| White | 3,110 | 0.09 (0.08 to 0.10) | 0.43 (0.40 to 0.47) | - | 4.69 (4.33 to 5.08) |
| Asian | 57 | 0.04 (0.03 to 0.06) | 0.16 (0.11 to 0.23) | 0.381 | 3.71 (2.21 to 6.24) |
| Black | 54 | 0.04 (0.02 to 0.06) | 0.26 (0.19 to 0.36) | 0.225 | 6.76 (3.76 to 12.2) |
| Other | 12 | 0.07 (0.03 to 0.16) | 0.21 (0.09 to 0.46) | 0.426 | 2.96 (0.95 to 9.20) |
| Mixed | 12 | 0.04 (0.01 to 0.12) | 0.31 (0.16 to 0.59) | 0.431 | 7.95 (2.14 to 29.5) |
| **Oesophageal-gastric** | | | | | |
|  | *n* | Normal test result incidence, % (95% CI) | High test result incidence, % (95% CI) | Interaction p-value | Odds ratio (95% CI) |
| White | 557 | 0.11 (0.10 to 0.12) | 0.21 (0.20 to 0.23) | - | 1.95 (1.73 to 2.21) |
| Asian | 11 | 0.06 (0.03 to 0.09) | 0.08 (0.03 to 0.14) | 0.393 | 1.38 (0.63 to 3.04) |
| Black | 7 | 0.09 (0.05 to 0.14) | 0.18 (0.09 to 0.28) | 0.987 | 1.94 (0.98 to 3.85) |
| Other | 1 | 0.05 (0 to 0.11) | . | . | . |
| Mixed | 1 | . | 0.38 (0.08 to 0.69) | . | . |
| **Uterine** | | | | | |
|  | *n* | Normal test result incidence, % (95% CI) | High test result incidence, % (95% CI) | Interaction p-value | Odds ratio (95% CI) |
| White | 302 | 0.03 (0.03 to 0.04) | 0.05 (0.04 to 0.07) | - | 1.49 (1.18 to 1.89) |
| Asian | 22 | 0.03 (0.02 to 0.06) | 0.09 (0.05 to 0.18) | 0.141 | 2.86 (1.24 to 6.62) |
| Black | 17 | 0.04 (0.02 to 0.08) | 0.07 (0.03 to 0.14) | 0.851 | 1.64 (0.62 to 4.32) |
| Other | 4 | 0.04 (0.01 to 0.17) | 0.11 (0.03 to 0.43) | 0.611 | 2.49 (0.35 to 17.8) |
| Mixed | 4 | 0.06 (0.02 to 0.20) | 0.06 (0.01 to 0.41) | 0.656 | 0.89 (0.09 to 8.56) |
| **Prostate** | | | | | |
|  | *n* | Normal test result incidence, % (95% CI) | High test result incidence, % (95% CI) | Interaction p-value | Odds ratio (95% CI) |
| White | 2,464 | 0.37 (0.33 to 0.40) | 0.44 (0.39 to 0.48) | - | 1.19 (1.10 to 1.29) |
| Asian | 44 | 0.13 (0.09 to 0.18) | 0.14 (0.08 to 0.26) | 0.877 | 1.13 (0.58 to 2.20) |
| Black | 147 | 0.76 (0.62 to 0.94) | 0.85 (0.62 to 1.16) | 0.721 | 1.11 (0.78 to 1.60) |
| Other | 28 | 0.71 (0.46 to 1.09) | 0.71 (0.33 to 1.50) | 0.703 | 1.01 (0.42 to 2.39) |
| Mixed | 12 | 0.22 (0.10 to 0.46) | 0.40 (0.17 to 0.97) | 0.461 | 1.84 (0.58 to 5.84) |
| **Breast** | | | | | |
|  | *n* | Normal test result incidence, % (95% CI) | High test result incidence, % (95% CI) | Interaction p-value | Odds ratio (95% CI) |
| White | 1,453 | 0.29 (0.27 to 0.31) | 0.38 (0.35 to 0.42) | - | 1.33 (1.19 to 1.49) |
| Asian | 63 | 0.21 (0.16 to 0.29) | 0.25 (0.16 to 0.41) | 0.706 | 1.20 (0.68 to 2.09) |
| Black | 49 | 0.22 (0.16 to 0.31) | 0.35 (0.22 to 0.55) | 0.595 | 1.57 (0.88 to 2.79) |
| Other | 9 | 0.22 (0.10 to 0.45) | 0.19 (0.05 to 0.76) | 0.611 | 0.89 (0.18 to 4.27) |
| Mixed | 10 | 0.29 (0.15 to 0.57) | 0.10 (0.01 to 0.74) | 0.209 | 0.35 (0.04 to 2.80) |
| **Lymphoma** | | | | | |
|  | *n* | Normal test result incidence, % (95% CI) | High test result incidence, % (95% CI) | Interaction p-value | Odds ratio (95% CI) |
| White | 1,297 | 0.09 (0.08 to 0.10) | 0.30 (0.28 to 0.33) | - | 3.49 (3.11 to 3.91) |
| Asian | 60 | 0.07 (0.05 to 0.10) | 0.23 (0.16 to 0.34) | 0.960 | 3.44 (2.07 to 5.72) |
| Black | 38 | 0.07 (0.05 to 0.12) | 0.23 (0.15 to 0.36) | 0.755 | 3.15 (1.66 to 5.95) |
| Other | 7 | 0.05 (0.02 to 0.15) | 0.23 (0.09 to 0.61) | 0.671 | 4.83 (1.08 to 21.6) |
| Mixed | 8 | 0.06 (0.02 to 0.17) | 0.21 (0.08 to 0.56) | 0.959 | 3.36 (0.84 to 13.5) |
| **Pancreas** | | | | | |
|  | *n* | Normal test result incidence, % (95% CI) | High test result incidence, % (95% CI) | Interaction p-value | Odds ratio (95% CI) |
| White | 1,022 | 0.05 (0.05 to 0.06) | 0.18 (0.16 to 0.20) | - | 3.37 (2.96 to 3.83) |
| Asian | 31 | 0.03 (0.02 to 0.05) | 0.10 (0.06 to 0.17) | 0.937 | 3.46 (1.71 to 7.01) |
| Black | 31 | 0.05 (0.03 to 0.08) | 0.16 (0.10 to 0.26) | 0.911 | 3.51 (1.73 to 7.12) |
| Other | 4 | 0.03 (0.01 to 0.12) | 0.10 (0.02 to 0.39) | 0.942 | 3.13 (0.44 to 22.3) |
| Mixed | 5 | 0.05 (0.02 to 0.15) | 0.09 (0.02 to 0.36) | 0.536 | 1.91 (0.32 to 11.5) |
| **Leukaemia** | | | | | |
|  | *n* | Normal test result incidence, % (95% CI) | High test result incidence, % (95% CI) | Interaction p-value | Odds ratio (95% CI) |
| White | 660 | 0.07 (0.00 to 0.08) | 0.14 (0.01 to 0.16) | - | 2.10 (1.79 to 2.45) |
| Asian | 14 | 0.03 (0.01 to 0.04) | 0.06 (0.02 to 0.11) | 0.918 | 2.21 (0.79 to 6.23) |
| Black | 17 | 0.04 (0.01 to 0.07) | 0.11 (0.04 to 0.18) | 0.673 | 2.58 (0.99 to 6.69) |
| Other | 0 | . | . | . | . |
| Mixed | 5 | 0.06 (0.04 to 0.13) | 0.12 (0.09 to 0.29) | 0.958 | 2.00 (0.33 to 12.0) |
| **Bladder** | | | | | |
|  | *n* | Normal test result incidence, % (95% CI) | High test result incidence, % (95% CI) | Interaction p-value | Odds ratio (95% CI) |
| White | 570 | 0.06 (0.06 to 0.07) | 0.10 (0.09 to 0.12) | - | 1.62 (1.37 to 1.92) |
| Asian | 8 | 0.02 (0.00 to 0.03) | 0.03 (0 to 0.06) | 0.951 | 1.69 (0.40 to 7.09) |
| Black | 4 | 0.01 (0 to 0.01) | 0.04 (0 to 0.08) | 0.173 | 7.84 (0.82 to 75.5) |
| Other | 4 | 0.05 (0 to 0.13) | 0.15 (0 to 0.35) | 0.584 | 2.81 (0.39 to 20.0) |
| Mixed | 2 | 0.13 (0 to 0.31) | . | . | . |
| **Kidney** | | | | | |
|  | *n* | Normal test result incidence, % (95% CI) | High test result incidence, % (95% CI) | Interaction p-value | Odds ratio (95% CI) |
| White | 666 | 0.04 (0.04 to 0.05) | 0.15 (0.14 to 0.17) | - | 3.60 (3.06 to 4.23) |
| Asian | 21 | 0.03 (0.02 to 0.05) | 0.07 (0.04 to 0.14) | 0.317 | 2.31 (0.99 to 5.42) |
| Black | 16 | 0.01 (0.01 to 0.04) | 0.13 (0.07 to 0.23) | 0.133 | 8.64 (2.78 to 26.8) |
| Other | 4 | 0.03 (0.01 to 0.13) | 0.11 (0.03 to 0.44) | 0.920 | 3.25 (0.46 to 23.1) |
| Mixed | 3 | 0.03 (0.01 to 0.13) | 0.05 (0.01 to 0.37) | 0.490 | 1.54 (0.14 to 17.3) |
| **Ovarian** | | | | | |
|  | *n* | Normal test result incidence, % (95% CI) | High test result incidence, % (95% CI) | Interaction p-value | Odds ratio (95% CI) |
| White | 542 | 0.05 (0.04 to 0.06) | 0.26 (0.23 to 0.30) | - | 5.48 (4.56 to 6.59) |
| Asian | 23 | 0.04 (0.02 to 0.07) | 0.17 (0.10 to 0.30) | 0.797 | 4.91 (2.15 to 11.2) |
| Black | 9 | 0.02 (0.01 to 0.06) | 0.09 (0.04 to 0.21) | 0.612 | 3.89 (1.04 to 14.5) |
| Other | 7 | 0.05 (0.01 to 0.20) | 0.44 (0.18 to 1.07) | 0.550 | 9.07 (1.76 to 46.9) |
| Mixed | 3 | 0.05 (0.01 to 0.20) | 0.09 (0.01 to 0.63) | 0.359 | 1.78 (0.16 to 19.6) |
| **Myeloma** | | | | | |
|  | *n* | Normal test result incidence, % (95% CI) | High test result incidence, % (95% CI) | Interaction p-value | Odds ratio (95% CI) |
| White | 539 | 0.06 (0.05 to 0.07) | 0.10 (0.09 to 0.11) | - | 1.62 (1.36 to 1.93) |
| Asian | 25 | 0.05 (0.03 to 0.08) | 0.08 (0.03 to 0.13) | 0.884 | 1.52 (0.65 to 3.52) |
| Black | 37 | 0.13 (0.08 to 0.19) | 0.18 (0.08 to 0.28) | 0.575 | 1.32 (0.67 to 2.63) |
| Other | 5 | 0.07 (0 to 0.15) | 0.15 (0 to 0.35) | 0.780 | 2.09 (0.35 to 12.5) |
| Mixed | 4 | 0.05 (0 to 0.11) | 0.13 (0 to 0.31) | 0.570 | 2.83 (0.40 to 20.4) |
| **Oral** | | | | | |
|  | *n* | Normal test result incidence, % (95% CI) | High test result incidence, % (95% CI) | Interaction p-value | Odds ratio (95% CI) |
| White | 411 | 0.05 (0.05 to 0.06) | 0.07 (0.06 to 0.08) | - | 1.35 (1.10 to 1.66) |
| Asian | 21 | 0.03 (0.02 to 0.05) | 0.05 (0.01 to 0.09) | 0.861 | 1.47 (0.57 to 3.81) |
| Black | 6 | 0.01 (0 to 0.02) | 0.03 (0 to 0.07) | 0.305 | 3.15 (0.63 to 15.6) |
| Other | 2 | 0.03 (0 to 0.08) |  | . | . |
| Mixed | 3 |  | 0.17 (0 to 0.35) | . | . |
| **Head and Neck** | | | | | |
|  | *n* | Normal test result incidence, % (95% CI) | High test result incidence, % (95% CI) | Interaction p-value | Odds ratio (95% CI) |
| White | 240 | 0.03 (0.03 to 0.04) | 0.03 (0.03 to 0.04) | - | 0.96 (0.72 to 1.28) |
| Asian | 17 | 0.03 (0.01 to 0.05) | 0.05 (0.01 to 0.09) | 0.426 | 1.49 (0.52 to 4.24) |
| Black | 10 | 0.02 (0.00 to 0.04) | 0.07 (0.01 to 0.12) | 0.056 | 2.99 (0.96 to 9.30) |
| Other | 3 | 0.05 (-0.01 to 0.11) | 0.06 (-0.05 to 0.17) | 0.874 | 1.16 (0.12 to 11.1) |
| Mixed | 0 | . | . | . | . |

Supplementary table X: adjusted incidence rates by cancer site and ethnicity.

Patients with a missing stage code were not included in the advanced stage analysis.
