## Supplementary materials 8 for "The impact of patient ethnicity on cancer incidence following platelet count and C-reactive protein tests in English primary care: a cohort study of 5 million patients"

Supplementary materials 8: Unadjusted one-year cancer incidence rates

| **Platelet cohort** | | | | | | |
| --- | --- | --- | --- | --- | --- | --- |
|  | **All cancer incidence, % (95% CI)** | **Advanced stage incidence, % (95% CI)*** | **Colorectal cancer incidence, % (95% CI)** | **Lung cancer incidence, % (95% CI)** | **Oesophageal cancer incidence, % (95% CI)** | **Uterine cancer incidence, % (95% CI)^+^** |
| **White** | 2.0  (1.9-2.0) | 56.9  (56.5-57.3) | 0.29  (0.28-0.29) | 0.25  (0.25-0.26) | 0.11  (0.10-0.11) | 0.08  (0.07-0.08) |
| **Asian** | 0.67  (0.64-0.70) | 49.4  (46.6-52.3) | 0.07  (0.06-0.08) | 0.05  (0.05-0.06) | 0.03  (0.02-0.04) | 0.05  (0.04-0.07) |
| **Black** | 1.0  (1.0-1.1) | 50.5  (47.5-53.4) | 0.10  (0.08-0.11) | 0.07  (0.06-0.08) | 0.04  (0.03-0.05) | 0.05  (0.03-0.06) |
| **Other** | 1.0  (0.9-1.1) | 53.7  (47.7-60.0) | 0.10  (0.07-0.14) | 0.10  (0.07-0.14) | 0.05  (0.03-0.07) | 0.03  (0.01-0.06) |
| **Mixed** | 1.0  (0.9-1.1) | 59.6  (54.0-65.0) | 0.13  (0.10-0.17) | 0.09  (0.07-0.12) | 0.07  (0.05-0.10) | 0.04  (0.02-0.07) |
| **CRP cohort** | | | | | | |
|  | **All cancer incidence, % (95% CI)** | **Advanced stage incidence, % (95% CI)*** | **Colorectal cancer incidence, % (95% CI)** | **Lung cancer incidence, % (95% CI)** | **Oesophageal cancer incidence, % (95% CI)** | **Uterine cancer incidence, % (95% CI)^+^** |
| **White** | 2.9  (2.9-3.0) | 59.5  (58.7-60.3) | 0.46  (0.44-0.47) | 0.44  (0.42-0.46) | 0.15  (0.14-0.16) | 0.07  (0.07-0.08) |
| **Asian** | 1.2  (1.1-1.3) | 52.9  (47.7-58.0) | 0.15  (0.12-0.18) | 0.11  (0.09-0.14) | 0.06  (0.04-0.08) | 0.07  (0.05-0.11) |
| **Black** | 1.6  (1.5-1.8) | 60.4  (55.5-65.3) | 0.19  (0.15-0.24) | 0.15  (0.11-0.19) | 0.09  (0.06-0.13) | 0.07  (0.04-0.12) |
| **Other** | 1.5  (1.3-1.8) | 51.2  (39.9-62.4) | 0.14  (0.07-0.24) | 0.15  (0.08-0.26) | 0.02  (0.003-0.09) | 0.08  (0.02-0.21) |
| **Mixed** | 1.3  (1.1-1.6) | 52.2  (39.8-64.4) | 0.13  (0.06-0.23 | 0.15  (0.08-0.26) | 0.08  (0.03-0.16) | 0.09  (0.02-0.23) |

**Supplementary : Unadjusted one-year cancer incidence rates from blood test date**

Figures indicate unadjusted one-year cancer incidence from blood test date, regardless of result, and reported as percentages with 95% confidence intervals.

* Percentage is reported from the cohort of patients diagnosed with cancer

^+^ Uterine cancer figures are for females only
